# Adaptive doctors: preparing tomorrow’s doctors for practice in the Anthropocene Epoch

**DOI:** 10.1101/2021.06.08.21258597

**Authors:** Claudia Slimings, Emily Sisson, Connor Larson, Devin Bowles, Rafat Hussain

## Abstract

**Background:** The future health workforce needs to be equipped with the knowledge, skills, and motivation to deliver sustainable healthcare and promote planetary health. The aim of this study was to design, implement and evaluate a new suite of planetary health learning activities piloted by medical students for a range of medical professionals.

**Methods:** The study consisted of three components: curriculum mapping, development of learning activities and evaluation. Curriculum mapping involved searching program learning outcomes using relevant search terms. Two learning activities were co-developed with medical students comprising of an e-learning component and an inquiry-based small group workshop presented to 99 2^nd^ year students. Evaluation consisted of pre- and post-learning knowledge quizzes and a student feedback survey.

**Results:** A total of 30 learning outcomes were identified with the majority located in the first two years of the four-year program. The overall evaluation response rate was 49.5%, and 19% completed the feedback survey. The mean pre- and post-lesson scores, respectively, were 7.09 (SD=1.84) and 9.53 (SD=1.69) out of a possible score of 12, increasing by 2.37 points on average (95% confidence interval [CI] 1.66-3.09). Overall, the new activities were rated as excellent/good by 84.2% of respondents. The e-learning module rated more highly as a meaningful learning experience than the workshop (89% v. 63.2%). The most common criticism was the length of time it took to complete the e-learning.

**Conclusion:** Students already had a good understanding of planetary health ‘facts’ and the e-learning lesson served to confirm, review and update their knowledge. Students embraced the opportunity to engage in interactive learning through the problem-solving group work activity. There is very little vertical alignment of environmental and climate issues across all four years of the medical program in our institution and a variety of learning approaches should be considered when revising the curriculum.

## Introduction

The unprecedented levels of harm to the environment from escalating human pressure is hazardous for human health, necessitating a “planetary health” approach to promoting human health and the health of the planet.^1, 2^ The World Health Organization has declared climate change the biggest health threat in the 21^st^ Century.^3^ The health care sector is vulnerable to the effects of, and contributes to, climate change, accounting for between 1 to 5% of total global carbon emissions.^4^ Australia has one of the highest levels of health care emissions in the world.^5^ Health systems need to work towards environmental sustainability, through lower carbon treatments, removing unnecessary procedures, preventing illness and promoting health.^6, 7^

While health professionals are essential for advancing environmentally sustainable health care and strengthening health systems for adaptation, they may also work to promote patient and community understanding, build community resilience, support education and research, and advocate for sustainable government policies.^8-11^ The Australian Medical Association (AMA) and their international counterparts – including the British Medical Association and the American Medical Association - have declared climate change a health emergency.^12^ The Position Statements of relevant peak bodies including the AMA and the Royal Australasian College of Physicians indicate the need to mitigate climate change, in order to prevent the health impacts of climate change, and adaptation in order to protect the health of vulnerable population groups.^13, 14^

The future health workforce needs to be equipped with the knowledge, skills and motivation to deliver sustainable healthcare and promote planetary health.^15-17^ The emergence of the SARS-Cov-2 virus (COVID-19) in 2019 has highlighted the need for medical education to provide an understanding of the “climate pandemic” and the interconnection between the health of people, animals and the environment (One Health).^18, 19^ However, there is currently a gap in formal learning opportunities in medical schools. In the USA, 56% of medical schools surveyed included some One Health subject matter.^20^ A recent student-led survey reported only 15% of medical schools globally included climate change and health in the curriculum, with student-led activities in a further 12%.^21^ The Planetary Health Report Card provides a detailed inspection of curricula, research, advocacy, support for student-led initiatives, and campus sustainability offered by medicals schools of the USA, Canada, UK, and Ireland.^22^ In the most recent report, no schools received an “A” for curriculum; 21% received a “B’, 48% a “C”, and 31% “D” or “F”.^23^.

Medical students are leading the development of learning resources to fill the current void.^24-27^ While peak bodies such as the Medical Deans of Australia and New Zealand (MDANZ) have developed and disseminated updated graduate outcome statements and learning outcomes for medical schools in Australia, these are yet to be formally incorporated into accreditation standards.^28^ Therefore the extent to which planetary health is included in medical school curricula is reliant on the motivation of individual educators who have to overcome barriers such as lack of curriculum space, resourcing, and institutional priorities.^29^ The inclusion of an indicator to the MJA-Lancet Countdown to track the incorporation of health and climate change education within Australia’s medical schools should drive progress.^30, 31^.

In response to the urgent need to improve planetary health offerings in medical education, faculty worked with students enrolled in the postgraduate medical program (MChD) at the Australian National University (ANU) to develop and implement new blended-learning activities. In addition to providing an opportunity for medical students to improve their planetary health knowledge, the project aimed to enhance student awareness of their future responsibility as health care professionals to promote and advocate for planetary health and sustainable health care. This paper reports on the design, implementation and evaluation of the new resources.

## Methods

### Curriculum mapping

The ANU Medical School curriculum map database was searched using the terms “climate”, “environment”, “planet*”, “sustain*”, “carbon”, “ecolog*”, “ecosystem”, “ecosocial” and “one health”. The term “advoc*” was also included to identify learning outcomes related to advocacy as this was an element included in the learning resource development. Previous research has indicated that medical curricula lack explicit learning outcomes and “hands-on” learning opportunities in health advocacy.^32^

The database predominantly includes level 1-3 learning outcomes for the MChD program provided in approved documentation. Level one includes the overarching program outcomes, level two the course level outcomes, level three the block level outcomes. Level four outcomes that describe the objectives of individual teaching sessions were identified through educational materials placed on the learning management system (LMS). Data were collected using Microsoft Excel, recording the number of learning outcomes within which search terms appeared in the curriculum and where they were located in the curriculum.

### Development and implementation of learning resources

A blended-learning approach was used to develop new learning activities drawing on constructivist learning theory.^33^ Two activities were developed comprising an e-learning module for transfer of core knowledge, and inquiry-based group work for development of more complex problem-solving skills. The group work component originally intended students to develop a ’pitch‘ for a strategy to advocate for planetary health issues to policy makers. Due to COVID-19 impacts on clinical teaching during 2020, this component could not go ahead in full and was substituted with a workshop. The activities were delivered to 2^nd^ year medical students over a three-week period in 2020 (e-learning module: weeks 1-2; group work: week 3) The development of the activities was led by two medical students employed as research assistants.

The e-learning module was created using Rise 360 (Articulate Global Inc.) and published on the LMS and will subsequently be made publically available. The learning outcomes were selected from proposed learning objectives developed by the MDANZ Climate Change and Health Working Group ^28^:

- Discuss the contribution of human activity to global and local environmental changes such as climate change, biodiversity loss and resource depletion.
- Describe the mechanisms by which human health is affected by environmental change.
- Explain how the health impacts of environmental change are distributed unequally within and between populations and the disparity between those most responsible and those most affected by change.
- Compare the carbon footprint of healthcare systems for different countries and describe the major contributors to each.
- Identify the role of health care professionals in advocating for policies and infrastructure that promote the availability, accessibility and uptake of healthy and environmentally sustainable behaviours.

The module comprised four parts. *Part 1 – Introduction*, provided an overview of planetary health. *Part 2 – The Problem:* the *Anthropocene and the Great Acceleration*, covered the contribution of human activity to environmental changes. *Part 3 – The Effect: how changes in the natural system are affecting human health*, described mechanisms by which human health is affected by climate change. *Part 4 – The Solution: Our role as future medical professionals*, discussed the role of health care professionals in advocating for policies and infrastructure that promote healthy and environmentally sustainable behaviours.

The inquiry-based component consisted of a 2-hour facilitated small group workshop designed to develop advocacy skills and identify practical ways to contribute to improving planetary health. The workshop was delivered using Zoom due to COVID-19 restrictions. An initial icebreaker activity asked students to discuss their experiences with advocacy in any setting. Students were then divided into small groups of four students (using the breakout room function) and selected a trigger scenario for the next activity. Scenario topics consisted of:

- designing an advocacy campaign to reduce the carbon footprint in the local hospital;
- assisting patients with continuity of care in the aftermath of bushfires;
- an advocacy campaign for an ecological intervention in the community;
- a campaign to lobby local government to implement policies to cut emissions produced by motorised transport;
- advocacy for healthy food options in medical practice and community settings to prevent illness and promote health.

Each scenario handout contained a brief outline, a task, suggested resources, and prompts. Students worked on the task in their group for 30 minutes and presented their findings back to the larger group. Facilitators were instructed to keep the students focused on the tasks and guide them through the process similar to the problem-based learning format.^34, 35^

### Evaluation

All 2^nd^ year medical students were invited to participate in the evaluation which consisted of pre- and post-learning knowledge quizzes and a feedback survey (Qualtrics, Provo, UT). Ethics approval was obtained from the ANU Human Research Ethics Committee (2020/448). An information session was held during the first week of the final teaching block of the year. Full details, instructions, and links to the quizzes and survey were provided on the LMS.

Students were instructed to complete the pre-lesson quiz before commencing the e-learning module. The total possible score for the quiz was 12. The post-lesson quiz consisted of the same questions used for the pre-lesson quiz and was made available to students, along with the feedback survey, at the end of the workshop. The Purdue Instructor Course Evaluation Service^36^ was used to guide question development for the feedback survey, covering domains of goals and objectives, relevance of content, teaching/learning of relationships and concepts, general student perceptions, student outcomes, diversity issues/respect/rapport, organisation and clarity. Three sets of questions were generated related to (i) the overall rating of “the course”, (ii) the e-learning, and (iii) the workshop.

Data were analysed using Stata V16 and figures generated in Excel. Descriptive analyses were undertaken using frequencies for categorical variables and mean and standard deviation (SD) or median and p25-p75 for numerical variables. Two sample t-test was used to compare the mean scores between different sub-groups, and a paired t-test to report the mean change in scores and 95% confidence intervals (CI) within groups. Free-text survey responses were coded for analysis using an inductive approach.

## Results

### Curriculum mapping

A total of 30 outcomes were identified of which five were level three outcomes and the rest were level four (Table 1). However, one level three outcome in year four on advocacy was not related to environmental or climate change. Of the 25 level four outcomes, 19 were located in phase one, of which 12 were provided by the population health theme, three by medical sciences, and two by professionalism and leadership. There was a lack of relevant learning outcomes in problem-based or case-based learning. *Environment* was the most common term included in learning outcomes, with fewer terms related to climate change and one that mentioned One Health.

**Table 1.** Curriculum mapping of learning outcomes related to environment, climate, sustainability, or advocacy.

| Level 3 learning outcomes |  |
| --- | --- |
| Phase 1 | Phase 2 |
| **Year 1:**<br>1. Describe how culture, society, socio-economic and *environmental* influences interact with biological factors to determine health and wellbeing.<br>2. Demonstrate skills in medical *advocacy*.<br>**Year 2:**<br>1. Analyse, evaluate and incorporate information from various sources to identify social, ethical, legal and human rights issues of presented medical cases in the context of global health issues including the ethical conduct of clinical trials, impact of trade agreements, pharmaceutical and insurance industries as well as new technologies (such as nanotechnology and artificial photosynthesis) on *sustainability* of a safe and healthy *environment* and health system quality and equity.<br>2. Evaluate and interpret the involvement of multiple body systems in complex diseases, including the interactions of biological determinants with the *environment*, society and culture, and apply this knowledge to clinical scenarios through effective communication, history taking and examination. | **Year 4:**<br>1. Articulate the need for *advocacy* in relation to the welfare of those with psychiatric and substance use disorders. |
| Level 4 learning outcomes |  |
| Phase 1 | Phase 2 |
| **Year 1:**<br>1. Describe the life-course, *One Health*, and Social Determinants of Health causal frameworks. (PH)<br>2. Define *environmental* health. (PH) | **Year 3:**<br>1. The genetic, *environmental*, and hormonal influences that determine normal growth. (CBL) |
| 3. Recognise the importance of *environmental* health as a health determinant. (PH)<br>4. List the major groups of *environmental* hazards. (PH)<br>5. Describe concepts of *environmental* exposure, including pathways, dose and effect. (PH)<br>6. Explain the relevance of *environmental* health to the doctor including history taking. (PH)<br>7. Identify Australian and international organisations engaged in *environmental* health advocacy. (PH)<br>8. Describe the primary drivers through which *climate change* will decrease human health. (PH)<br>9. Examine the methodological obstacles to understanding the effect of future *climate change* on health, and approaches to circumvent these obstacles. (PH)<br>10. Discuss some of the barriers to action on *climate change*. (PH)<br>11. Describe the individual, *climate* and *environmental* allergens that contribute to the causation of thunderstorm asthma epidemics. (PH)<br>12. Discuss the impact of demographic, migration, globalisation and *environmental* changes on global health. (PH)<br>13. Identify the factors that are associated with an increased risk of asthma, specifically genetic, associated medical conditions and *environmental* factors. (MS)<br>14. Appreciate the difference and importance of *environmental* and genetic factors that influence the drug response. (MS)<br>15. A grasp of the broader social, cultural and political role played by the healthcare professions in public life and in shaping the legal and regulatory environment (*advocating* for change). (PAL)<br>16. To understand the genetic/*environmental* contributions and common pathologies in | 2. Know principles of *advocating* for your own career. (FRS)<br>**Year 4:**<br>1. Describe and explain how a psychiatric presentation may be an interaction between genetic vulnerability and *environmental* (bio-psycho-social) factors/stressors. (PAM)<br>2. Understand the comprehensive *environment*, social, family and cultural elements to the maintenance of indigenous good mental health. (PAM)<br>3. Discuss these topics: Dental care, Travel including seat belt use and car safety, Occupation, Sex in pregnancy, Exercise and diet, *Environmental* hazards in pregnancy. (WH)<br>4. Develop skills for communication in roles including health *advocacy*. (PH) |
| abnormalities of the hypothalamo-pituitary-thyroid-adrenal axis (PBL)<br>**Year 2:**<br>17. Understand at a basic level ethical and human rights obligations of doctors to engage with issues of global health such as corporatogenic *climate change*. (PAL)<br>18. List key *environmental* aetiology factors associated with schizophrenia. (MS)<br>19. Discuss the genetic and *environmental* contributions to gallstone formation. (PBL) |  |

Abbreviations: PH – Population Health; MS – Medical Sciences; PAL - Professionalism and Leadership; PBL - Problem-based learning; Case-based learning; FRS – Fixed Resource Session; PAM – Psychiatry and Addiction Medicine; WH – Women’s Health.

### Evaluation

Out of 99 students enrolled in the course, 88 attempted the e-learning and 81 attended the workshop; 49 students participated in the evaluation study (response rate 49.5%). All 49 consented to the pre- and post-lesson knowledge tests, and 44 consented to the feedback survey.

### Knowledge tests

Most students in the study (n=46) completed the pre-lesson test, of which 44 students accessed the e-learning module. Eight students (18.2%) accessed the e-learning before even starting the quiz and 21 students (47.7%) accessed the e-learning after commencing but before completing the pre-lesson quiz (Supplementary Figure 1). The median time between pre-accessing the e-learning and completing the quiz was 1.16 minutes (p25-p75= 0.32-27.2 minutes).

The post-lesson test was completed by 34 students; 32 completed both pre- and post-lesson tests. The total number of correct answers ranged from 3 to 10 for the pre-lesson quiz and 6 to 12 for the post-lesson quiz. The mean pre- and post-lesson scores were 7.09 (SD=1.84) and 9.53 (SD=1.69), respectively, increasing by 2.37 points on average (95% confidence interval [CI] 1.66-3.09; p<0.0001) (Table 2).

**Table 2.** Pre- and post-lesson knowledge quiz results

|  | Pre-lesson score |  | Post-lesson score |  | Change in score |  |
| --- | --- | --- | --- | --- | --- | --- |
|  | n | Mean (sd) | n | Mean (sd) | n | Mean (95% CI) |
| Overall | 46 | 7.09 (1.84) | 34 | 9.53 (1.69) | 32 | 2.37 (1.66-3.09) |
| Accessed e-learning before starting pre-quiz |  |  |  |  |  |  |
| No | 36 | 7.31 (1.67) | 25 | 9.80 (1.53) | 25 | 2.32 (1.48-3.16) |
| Yes | 8 | 6.25 (2.43) | 7 | 8.86 (2.27) | 7 | 2.57 (0.81-4.33) |
| p-value* |  | 0.145 |  | 0.205 |  | 0.772 |
| Accessed e-learning before completing pre-quiz |  |  |  |  |  |  |
| No | 23 | 7.61 (1.47) | 15 | 10.0 (1.56) | 15 | 2.07 (0.99-3.14) |
| Yes | 21 | 6.57 (2.09) | 17 | 9.23 (1.82) | 17 | 2.65 (1.60-3.69) |
| p-value* |  | 0.062 |  | 0.215 |  | 0.417 |
| Completed e-learning |  |  |  |  |  |  |
| No | 18 | 6.78 (1.99) | 10 | 9.22 (1.99) | 9 | 2.89 (0.87-4.90) |
| Yes | 28 | 7.29 (1.74) | 24 | 9.67 (1.58) | 23 | 2.17 (1.44-2.91) |
| p-value* |  | 0.366 |  | 0.472 |  | 0.102 |

\*two sample t-test for difference in means between two groups

Both pre- and post-lesson scores were lower for the group that accessed the e-learning before either starting or completing the pre-lesson quiz than the group that did not, however the differences were not statistically significant, and there was no difference in the magnitude of change in knowledge between pre- and post-lesson tests (Table 1). Similarly, there was little difference in the scores or change in knowledge between those who completed the e-learning and those who did not.

### User feedback

The feedback survey was completed by 19 students (9 males, 8 females, 2 unspecified; age range 22-38 years). The overall course presentation was rated as excellent or good by 16 students (84.2%). Students generally agreed with all statements related to the new resources collectively, particularly the use of up to date materials and creative planning of the course (Figure 1).

**Figure 1.**
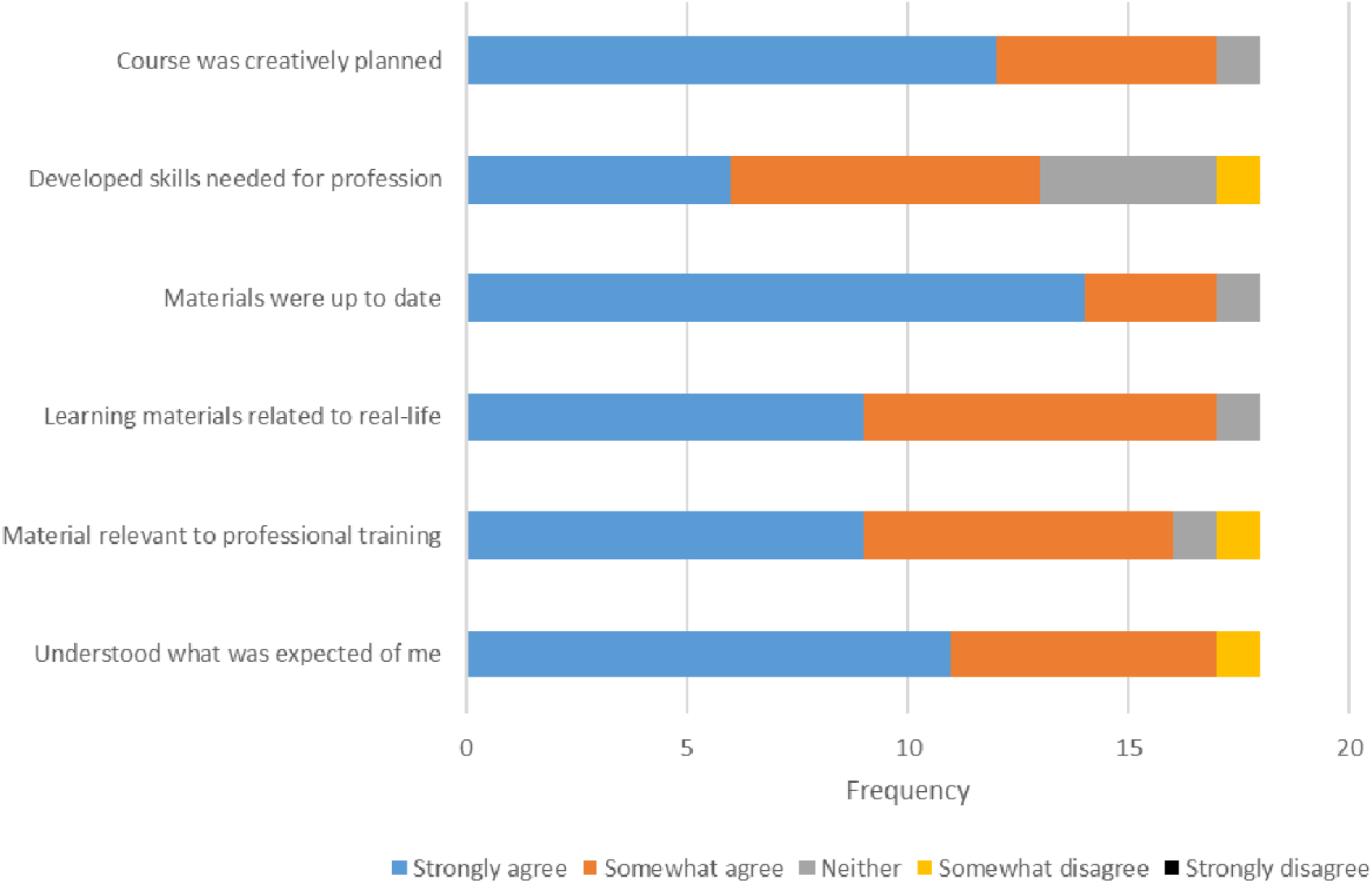
Frequency of responses to statements about the learning activities in general

The e-learning module itself was well rated, with all students who responded agreeing that the module made good use of examples and built an understanding of concepts (Figure 2 top panel). Most felt it provided a meaningful learning experience (n=17, 89%). The majority also felt that it improved their knowledge on climate change and health or that the module was consistent with their prior expectations (n=15, 78.9%).

**Figure 2.**
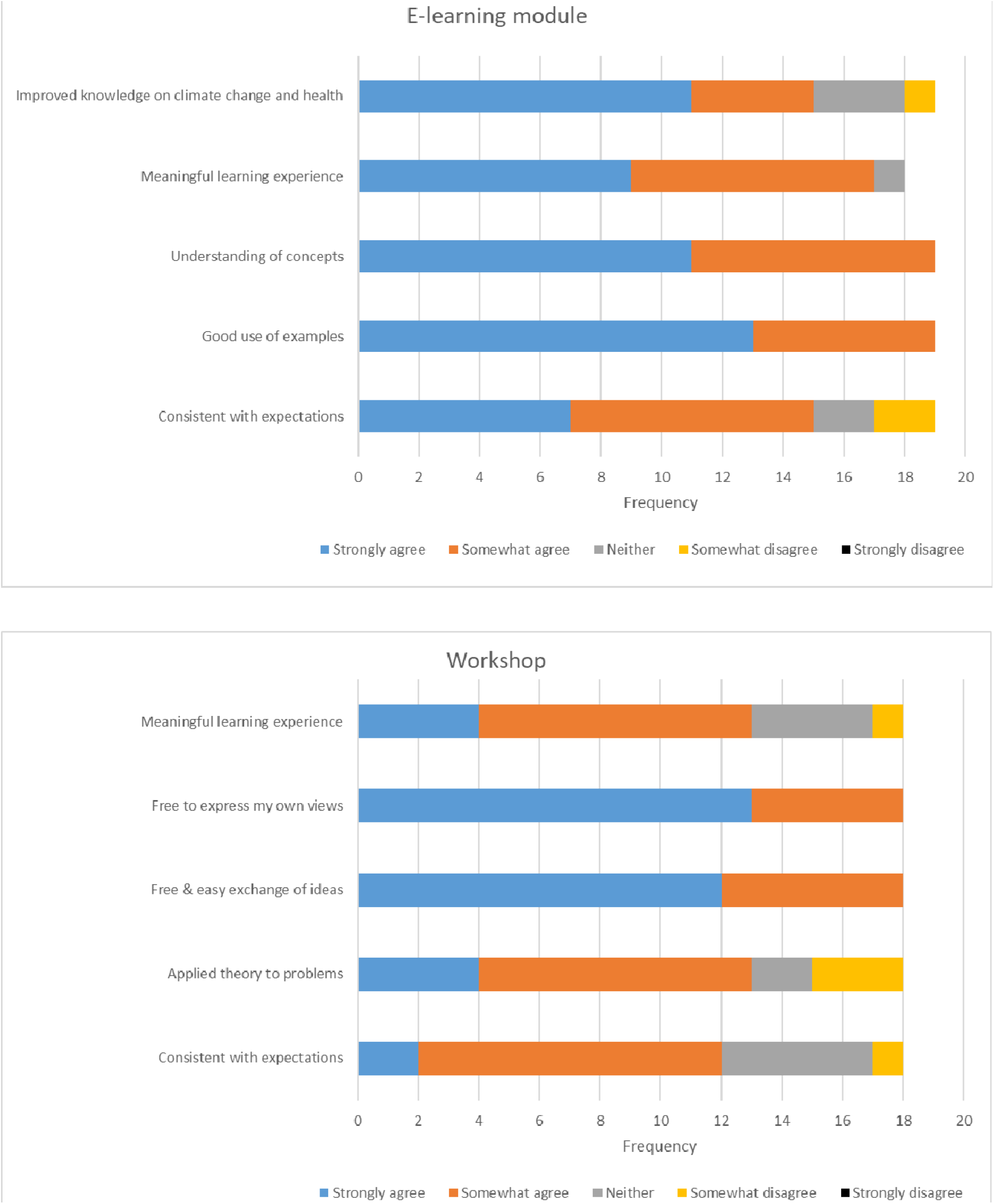
Frequency of responses to statements about the e-learning module and workshop

All students felt that they could express their views and there was free and easy exchange of ideas during the workshop (Figure 2 bottom panel). Fewer students (n=13, 68.4%) felt that the workshop was a meaningful learning experience or was consistent with their expectations (n=12, 63.2%) compared to the e-learning module. However, there were no instances of strong disagreement with any of the statements presented.

Fourteen students provided free text comments regarding the e-learning module from which 38 responses were identified and coded to nine response categories (Figure 3). The length of the e-learning module was most commonly commented on, accounting for 19% of responses (n=7) and all indicating that it was too long. Other responses regarding repetition (n=3) and content (n=4) provided insights for module improvement.

**Figure 3.**
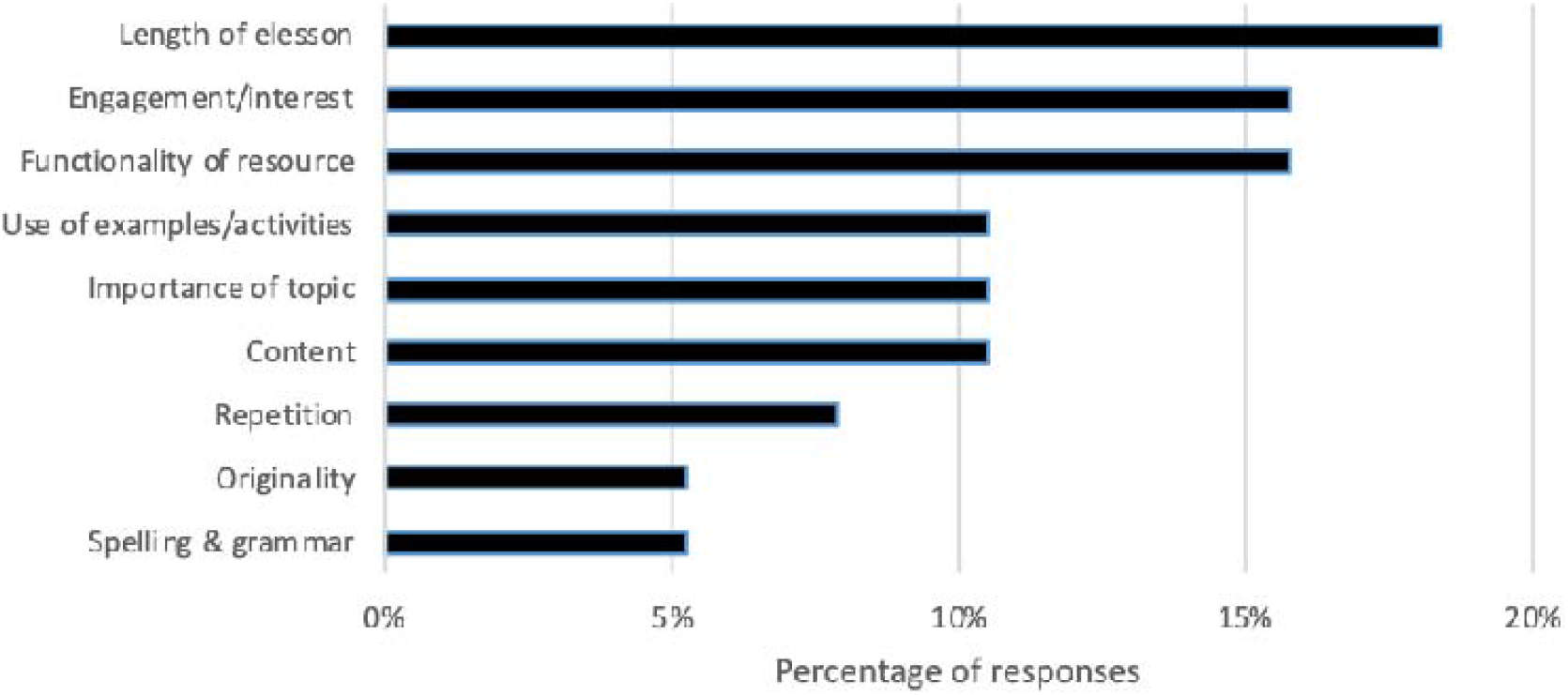
Categories identified from coding of full text responses about the e-learning module

> “The lesson was too long and highly repetitive, which made synthesising information from each section difficult. Took me multiple days and sittings to complete. However, prefer this platform over [usual platform] and appreciated the interactive nature of the module.”
>
> “Excellent resource, a little repetitive at times but that is my only criticism. I think this is a very important and previously undervalued part of the curriculum.”
>
> “Maybe some streamlining of the intro/conclusions and summaries could make it less repetitive?”
>
> “A lot of the content was common knowledge”.

Eight students commented on the workshop, from which 15 responses were coded to five categories: workshop format (5 responses), content (5 responses), interaction with peers (3 responses), content (1 response), and linkage between e-learning and workshop (1 comment).

> “I felt it was too focussed on advocacy”
>
> “I liked the interactive nature of the workshop. It was good to hear other people’s views.”
>
> “Less time could be allocated to the advocacy/icebreaker discussion and some time to go through the levels/steps of advocacy may have been beneficial. I quite enjoyed the activity in planning an approach to advocating on a topic and then presenting it back to the group.”
>
> “It might be more fun to have participants design an advocacy campaign poster or specific new initiative that they would like to implement instead of just giving a broad overview of the issue which most people already know about.”

Seven students provided responses to the course generally, of which more than half were very positive (n=4) demonstrating their enjoyment of the course, the relevancy of the topic and the mode of delivery. The less positive responses related to the length of the e-learning module with a suggestions to provide the course over a few interactive sessions.

## Discussion

This study reports the development, implementation and evaluation of new planetary health educational activities for a postgraduate medicine program in an Australian university. Medical professionals need to promote planetary health through delivering sustainable healthcare, managing climate-related illness, and local health promotion and prevention.^15-17^ Core competencies include identification of vulnerable patients, risk factor management, diagnosis and management of climate-related physical and mental illness.^16^ In addition, the shifting distribution of health needs with environmental change, plus the geographic variation of the impact of environmental change on communities, requires medical professionals to be literate in planetary and public health.^16^ There is currently little formal education on planetary health and sustainable healthcare in medical schools globally, however student bodies, medical education academics and planetary health scientists are driving curriculum reform.^15, 21, 28, 30, 37^

At our medical school we developed two new learning activities as a first step to improving the planetary health content in the curriculum. Engaging students actively in their learning, where they are ‘producers’ or ‘partners’ of knowledge, rather than consumers was a key feature of the process.^38, 39^ The current student-led initiatives to improve planetary health in medical programs highlights the importance of engaging with students to update contemporary curricula.^22, 26^ As the survey results showed, students already had a good understanding of climate-health ‘facts’ and the e-learning lesson served to confirm, review and update their knowledge. Students embraced the opportunity to engage in interactive learning through the problem-solving group work activity. One benefit of co-designing with students was being able to identify and test activities that would actively engage students. This resulted in the inclusion of clinical scenarios related to planetary health issues in each part, such as a patient with heat stroke and pollution-induced respiratory issues.

Student feedback indicated that the opportunity to design a campaign rather than workshopping existing ideas would be welcome. The project originally intended for students to develop a ‘pitch’ for a strategy to advocate for planetary health issues to policy makers for the group work component. Following a brain storming workshop, student groups were to be given one week to develop their pitch which would be peer-assessed formatively through student presentations the following week. Peer assessment has several advantages, such as encouraging students to critically reflect on each other’s work, be involved in the assessment process, providing more feedback than that obtained from just 1-2 educators, development of generic skills such as constructive feedback, critical appraisal, and working co-operatively.^40, 41^ Due to the impact of the COVID-19 pandemic on student clinical placements in 2020, this component could not proceed due to the prioritisation of timetable space to accommodate clinical skills catch-up teaching and was substituted with a single, non-mandatory workshop.

This study has several limitations. A low response rate to the feedback survey limits the representativeness of the findings, however, the consistency between responses on domains such as the time taken to complete the e-learning, balanced with the positive responses to the initiative generally, suggest the responses are valid and will be taken into consideration when revising the learning activities for future use. The disruption to teaching due to COVID-19 resulting in the implementation of a ‘plan b’ for the inquiry-based component was sub-optimal and highlighted the barriers to introducing new curricula into medical programs with already full curricula. There was some irony in the institutional de-prioritisation of new planetary health learning activities during a global pandemic. Curriculum mapping was limited to searching available sources of learning outcomes and may have missed some sources of level four outcomes.

While the new activities are welcome, they are only a small, initial step. A variety of learning approaches are available to medical educators that can be vertically integrated into the curriculum including problem- or case-based learning, clinical skills training, and placement projects and through assessment including OSCEs.^15, 16^ Students in our program have the opportunity to engage with placement projects through the Phase 2 Population Health project. Students can elect to undertake either prevention/evaluation/policy research, conduct a clinical audit, or develop a health promotion video. In each of these three streams there is potential for students to work on planetary health relevant topics, such as the health impacts of waste incineration conducted with the Public Health Association of Australia.^42^

Curriculum mapping identified minimal vertical alignment of environmental and climate issues across all four years of the medical program in our institution or through different learning activities. Therefore, a full curriculum review is required to constructively align learning outcomes, learning activities, assessment and feedback for students to be equipped with core competencies to promote planetary health. ^15^

In conclusion, health professionals are essential for promoting planetary health and providing sustainable healthcare. It is the responsibility of educational institutions to prepare students with the knowledge, skills and competencies to work in a world destabilised by declining planetary health. Medical students are willing to engage in this topic and medical schools are doing a disservice to the future health workforce by not addressing this.

## Data Availability

Data will be archived with the ANU Data Commons and available on application

---

**Supplementary Figure 1.**
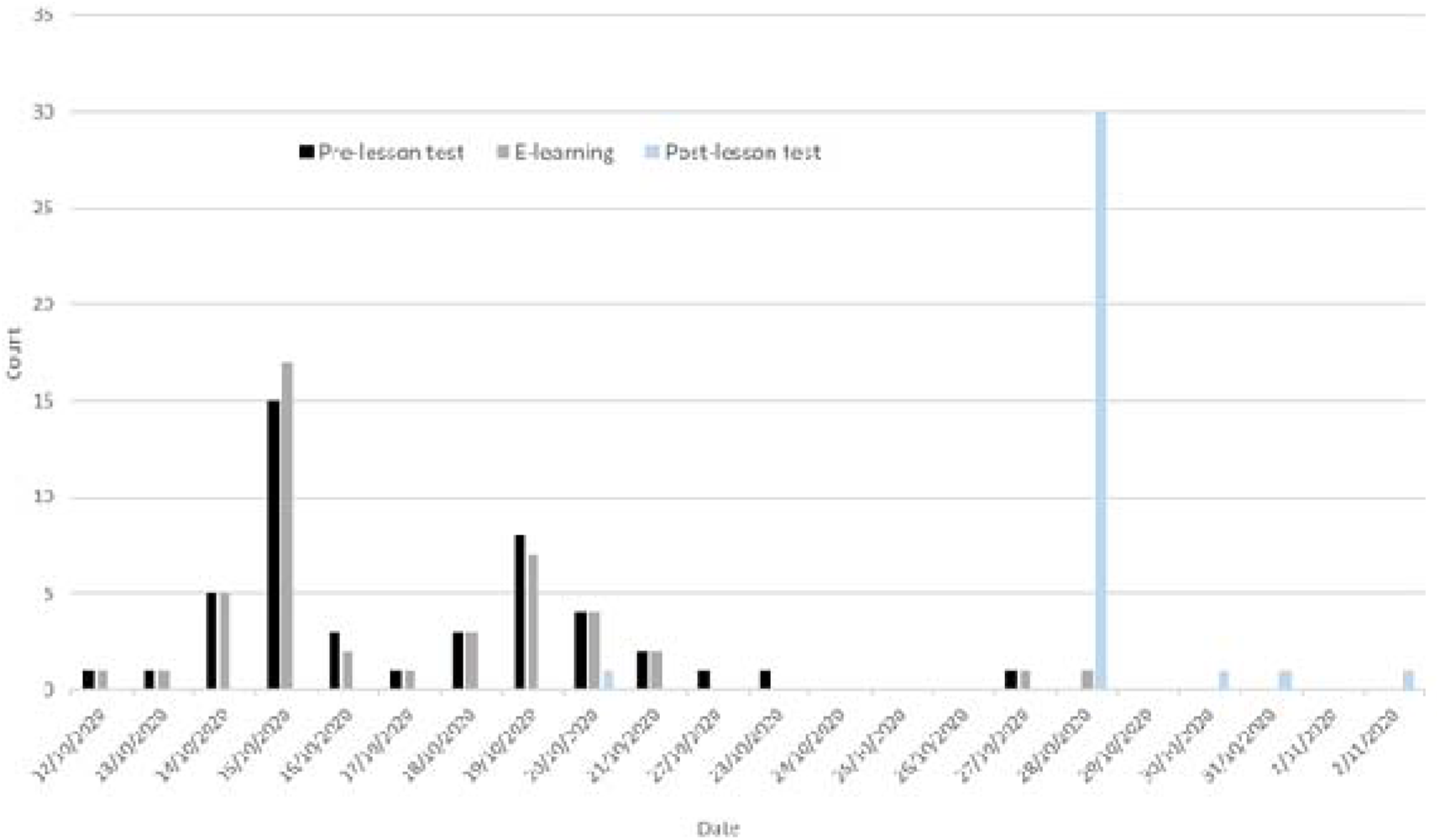
Dates students first accessed the E-learning module and completed the pre- and post-lesson knowledge tests

